# Forecasting the SARS-CoV-2 effective reproduction number using bulk contact data from mobile phones

**DOI:** 10.1101/2020.10.02.20188136

**Authors:** Sten Rüdiger, Stefan Konigorski, Jonathan Edelman, Detlef Zernick, Alexander Thieme, Christoph Lippert

**Affiliations:** NET CHECK GmbH, Berlin, Germany; Digital Health - Machine Learning, Hasso-Plattner-Institut, Universität Potsdam, Germany; Department of Radiation Oncology, Charité - Universitätsmedizin Berlin, Germany; Berlin Institute of Health (BIH), Berlin, Germany; Stanford Center for Biomedical Informatics Research, Stanford Medicine, Stanford, USA

## Abstract

Over the last months, cases of SARS-CoV-2 surged repeatedly in many countries and could often be controlled with non-pharmaceutical interventions such as social distancing. We analyzed de-identified GPS tracking data from 1.15 to 1.4 million cell phones in Germany per day between March-November 2020 to identify encounters between individuals and statistically evaluate large-scale contact behavior. Using graph sampling theory we estimated the contact index (CI), a metric for number and heterogeneity of contacts and found that the contact index, and not the total number of contacts, is an accurate predsictor for the effective reproduction number *R*. A high correlation between CI and *R* occurring more than two weeks later allows timely assessment of the social behavior well before the infections become detectable. The CI quantifies the role of superspreading and allows assigning risks to specific contact behavior. We provide a critical CI-value beyond which *R* is expected to rise above 1 and propose to use it to leverage the social distancing interventions for the coming months.

---

In December 2019, the novel coronavirus, SARS-CoV-2, caused a sustained pandemic with more than 60 million confirmed cases and more than 1.4 million deaths as of November 2020. SARS-CoV-2 is highly contagious with an estimated basic reproductive number *R*0 between 1.5-4 [1, 2, 3, 4]. It may spread invisibly in the community until a local outbreak becomes noticeable by a larger number of severe clinical cases. In the absence of a vaccine, non-pharmaceutical interventions (NPIs) have been considered an important tool to contain the spread of the virus. In Germany, before the implementation of strict interventions, an exponential growth of case numbers led to an effective reproduction number *R* of about 3. Beginning March 13, 2020, public events with more more than 50 visitors were banned, followed by a nationwide lockdown with a general ban on public contacts on March 23, 2020. From mid-March there was a steady decline of *R*, followed by a plateau of *R* around 1 and an average number of less than 1,000 new cases daily from May to the end of September. Sufficiently low levels of daily new infections allowed for the release of interventions. However, as SARS-CoV-2 could not be completely eradicated from the population, a second wave began in October, and new restrictions were introduced with interpersonal distance and rapid detection of local outbreaks remaining key.

Due to asymptomatic carriage of SARS-CoV-2 [5, 6, 7], a large portion of infected individuals remains undetected. Furthermore, it is estimated that 44% of secondary cases are infected through pre-symptomatic transmission events [8]. With an estimated incubation time of 5 days [3],a reported time of 6 days between symptoms and diagnosis based on laboratory tests [9], laboratory testing does not appear to be ideal for rapid outbreak detection given the short infection doubling time of SARS-Cov-2 of 1.4 to 2.5 days [10]. Thus, other methods are needed which allow for the early detection of outbreaks and forecast of the outbreaks’ intensity. Soon after the beginning of the pandemic, several cell phone apps have been developed to monitor the health of the population or record contacts between users based on device-mounted Bluetooth. While the technology is promising, the results in several countries, including Germany, have not met expectations [11].

In this study, we analyze position-derived encounters or “contacts” of cell phones. We hypothesize that individual location history data enabled by the GPS information of mobile phones can capture contact behavior of the population statistically and in this way allows for conclusions on the epidemic spread even if information on the infection states is not included. For our analysis we assume that each cell phone is used always by the same individual. For each cell phone in our representative and anonymous panel of more than one million devices we obtain records that contain up to several hundreds messages per day and device. We then project the positions for each message to a predefined tile of about 8m *×* 8m. Using an identification number of the tile we then scan for coincident presence of two different individuals on the same tile with the same time stamp rounded to two minutes, see Fig. 1A and Supplementary Material. For simplicity, we define as one contact a pair of individuals that encountered one or more times during one day. The collected data covers about 1 % of the German population and the entire period of the pandemic, including weeks before its beginning.

**Figure 1:**
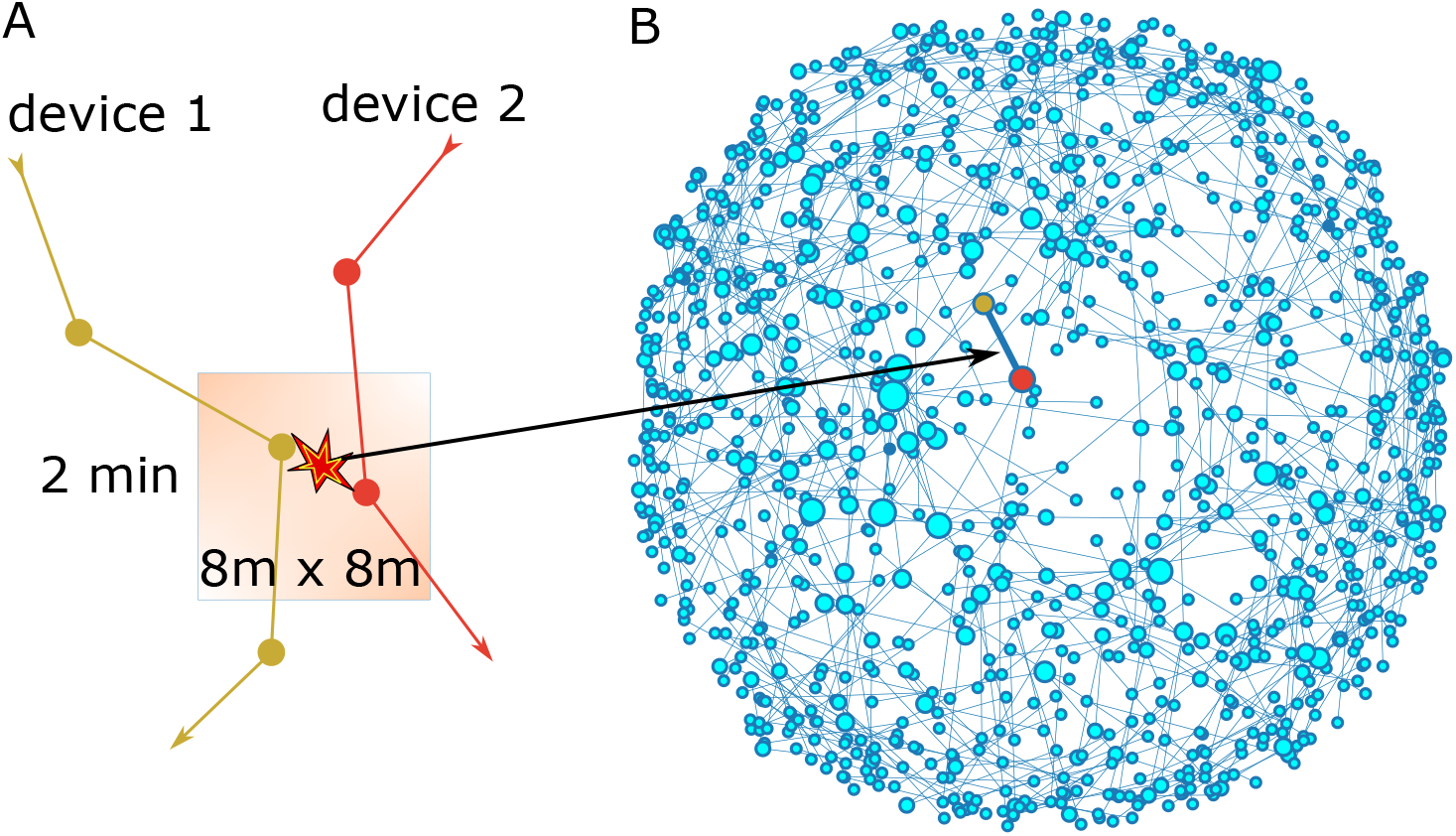
(A) Our method of identifying encounters rests on the co-location of two devices on a geolocation tile within a time interval of two minutes. In the contact graph pairs of devices are linked if a co-space-time-location was found at least once per day. The network shown in (B) is the subnetwork of cell phones located in Leipzig on Feb 29. The size of the dots corresponds to the degree, or number of contacts, of the node. The layout of nodes is obtained from a spring-force algorithm [12].

For a detailed analysis of the data we use methods from complex network science. We consider a graph of contacts for each day by assigning each device to a node of the network while each contact defines an edge between the respective nodes Fig. 1B. We then focus on the graph parameters that determine the simplest measures for epidemic modeling: the mean degree and the heterogeneity of the degree distribution. For a statistical estimation of the moments of the degree distribution for the underlying “real” Germany-wide network we use graph sampling theory described in the Supplementary Material.

In Fig. 2 we compare the evolution of the effective reproduction number (panel A) estimated from the Nowcast case numbers for Germany (seven-day-*R* value from [13]) with the evolution of the graph parameters. Fig. 2B shows that the mean number of contacts of individuals per day was visibly reduced in the mid of March. This holds particularly in the initial phase of the outbreak after which the weekly average number of contacts per person drops from around 20 to less than 7 (March 27). However, beginning in mid of April the number of contacts increases again and reaches a relatively stable plateau of about 75 % of the pre-outbreak level for the rest of the time. Interestingly, the contact number increases with the beginning of mask use suggesting that the perceived additional safety of the masks leads to a relaxation of personal distance.

**Figure 2:**
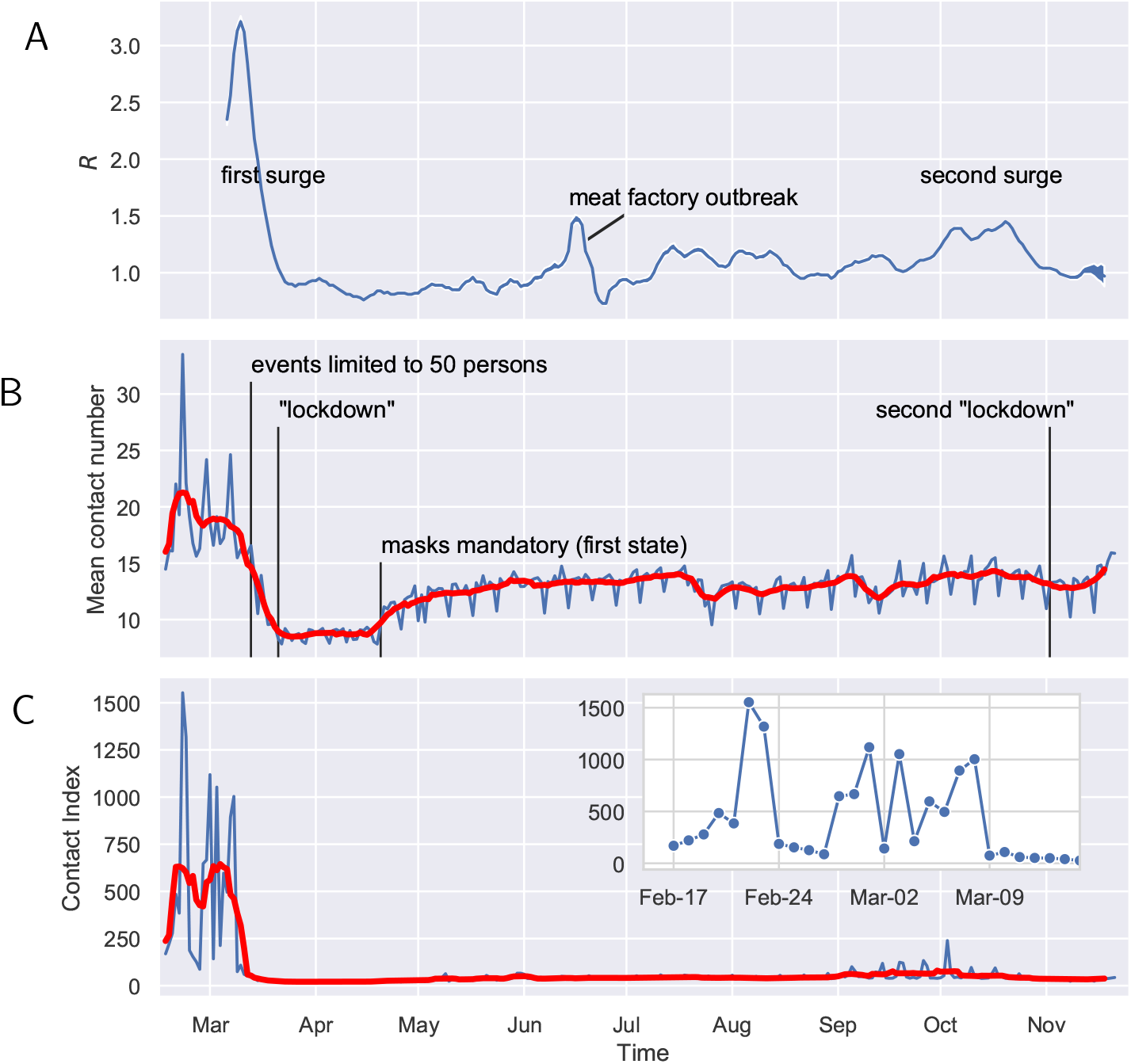
(A) The effective reproduction number *R* for Germany based on a summation of case numbers for seven days. (B) Mean number of contacts per day from cell phone records (blue) and seven day moving average (red). (C) Contact Index per day (blue) and seven day moving average (red).

The two curves, for *R* and the mean contact number, roughly correspond. However, we note that the *R* value decreases synchronously with the mean degree until both hit minimal levels, which is not intuitive since the contacts should precede the infection for a few days. As well, there is the substantial increase of the number of contacts after April which is not accompanied by an equally strong increase in *R*. While the number of contacts recovers 75% of its maximal value, *R* stays at values around 1.0 for much of the summer months. We thus conclude that the number of contacts does not sufficiently reflect and anticipate the subsequent evolution of *R*. For this, a more complex metric is needed.

It is known that for heterogeneous networks the reproduction number *R* on graphs generally follows the ratio of second to first moment of the degree distribution but not the first moment (or mean contact number) [14]. The degree distribution can be very broad in the case of social networks and the ratio gives a very different number than the mean (the numbers are equal if the contacts are homogeneous). Particularly we find this to be the case in our contact graph indicating a strong role of heterogeneity in the contact behavior. In the following, to distinguish the incidence-based *R* from the contact-based number, we introduce the “Contact Index” (CI) for the statistical estimate of its population value and compare it to *R* derived from the incidence numbers.

In Fig. 2C the curves show the evolution of the contact index (blue) and its seven-day moving average (red). After the first lockdown, the contact index stays relatively constant and remains low even after relaxation of lockdown and during the summer. This suggests that it reflects and anticipates better the evolution of *R* then the mean number of contacts. The inset shows the detailed evolution of the CI before the first lockdown and gives an idea how the “normal” social behavior of the population is reflected in the CI. First, a strongly increased CI, up to 1000 and more, is seen during weekends, particularly on Fridays and Saturdays. On other days of the week the CI is at a few hundreds. There are notable exceptions during the working days such as March 3, a Tuesday. On this day we identified a soccer game of the German cup to be largely responsible for the increase. The evolution of the Contact Index during the first wave and its relation to the reproduction number can be further inspected in Fig. 3A. Indeed, we can see that strong decreases followed a number of political interventions: cancellation of soccer games (three games in the first division on March 8, one final game on March 9) and other large public gatherings, limiting of public concerts etc. to a maximum of 50 persons (March 13), closing of school (March 16), social distancing measures (“lockdown”, March 23). It should, however, be kept in mind that companies shifted to home-office during this time. The specific effect of the various measures is thus hard to distinguish from self-imposed social distancing. Finally, we notice that the CI reflects the return of *R* towards 1 much better then the total number of contacts. The CI anticipates the *R* decrease by about one week during the first wave, while, as described above, the contact number falls concurrently with *R*.

**Figure 3:**
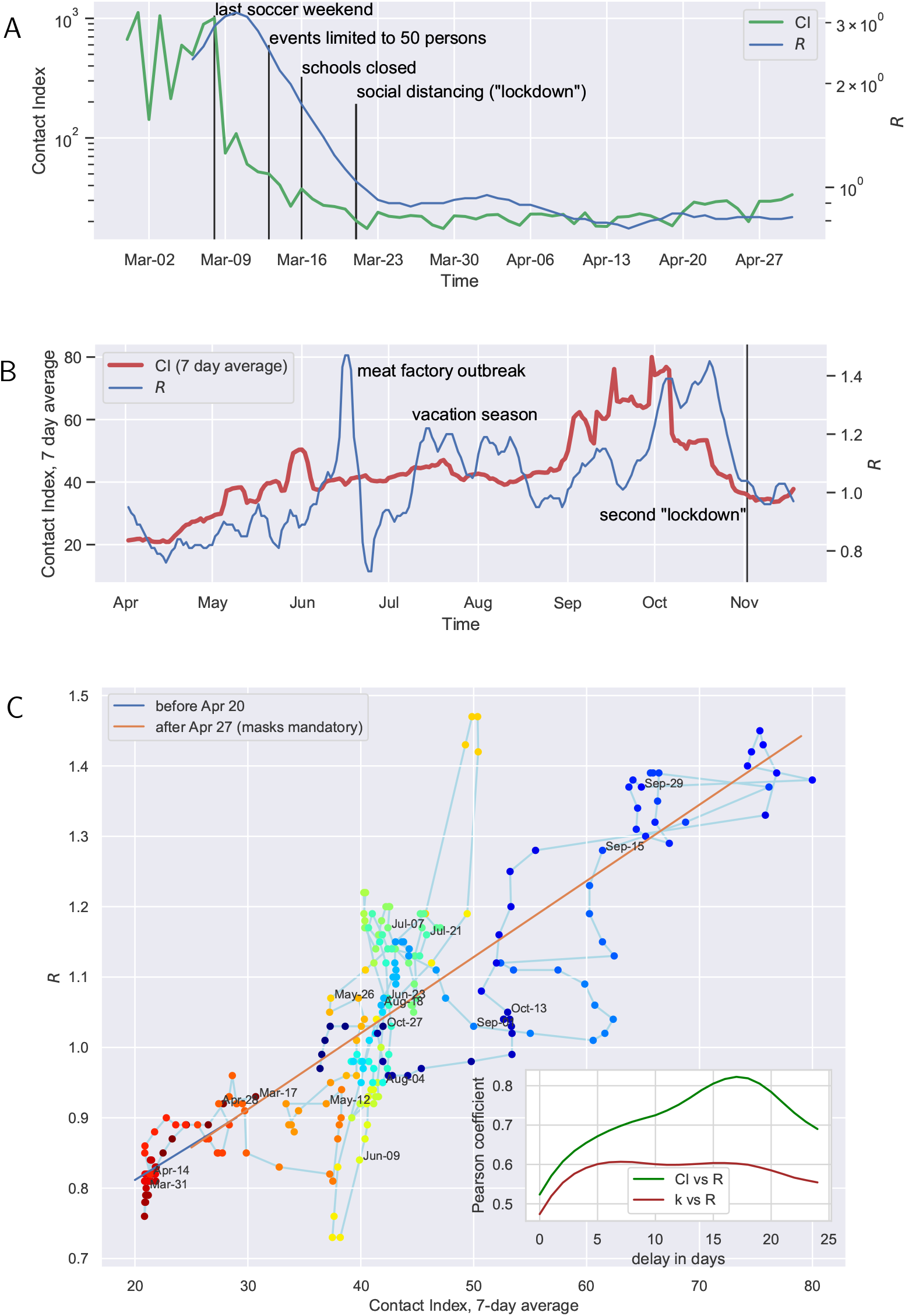
(A) The evolution of CI and *R* during and after the first surge. We have also added time points of notable political interventions (vertical black lines). (B) Evolution post first wave shows a substantial and related increase of *R* and Contact Index. (C) *R* and the Contact Index clearly exhibit an alm6ost linear relation if plotted with a shift of 17 days. Colors denote different weeks as indicated. The inset shows the Pearson correlation coefficient against the number of days in delay

The Contact Index also shows a strong association with the outbreak’s evolution in the phase after the first wave and during the second wave. Fig. 3B shows a zoom of the time evolution after the first wave, exhibiting a concurrent evolution of *R* and CI. Generally, the CI anticipates the infection numbers. This holds particularly for the onset and decay of the second surge in October, which is anticipated by a strong increase and decay of the CI. A notable omission from this co-evolution is the large but short effect of a local outbreak in June (with 1,413 infections [15]) as well as an increase of *R* for a few weeks in July and August. We assume that a part of these infections was due to travellers that returned from vacation in other countries with higher incidence rates. We can therefore not expect that a similar behavior is seen in the CI that only reflects connectivity within the country.

The correlation of CI and *R* can be assessed quantitatively from Fig. 3C which plots the *R* values versus the contact index at 17 days earlier showing the predictive power of the CI. The Pearson correlation coefficient is maximized for a delay of 17 days between the CI and *R* with a coefficient of 0.82 (see inset, 95 % confidence interval 0.78-0.86). A much smaller coefficient is obtained for the correlation of *R* and the mean contact number (red curve in the inset).

A linear regression of *R* and CI for a delay of 17 days was applied for two phases: one before the mandatory use of masks (blue diagonal line in 3B) and after masks were used widespread (red line). We did not find a substantial difference in the linear approximation. Thus, in contrast to studies that find a protective effect of mask wearing ([16] but also see [17]), our data does not provide significant evidence for an effect of masks in limiting of infection behavior. The linear regression model predicts that a contact index of CIcrit ≈ 38 (95 % confidence interval 19-57) or more would drive the infection behavior into a sustained super-critical regime with *R >* 1. This observation can be useful for the tuning of social distancing measures and the timely assessment of the measures.

It is instructive to analyze the causes for the large CI before the lockdown and the reduction of the CI after a lockdown. We found that indeed a very broad distributions of contact numbers for individuals is responsible for the large CI values before the lockdown. After each surge, this distribution becomes much more narrow leading to a smaller CI. This demonstrates the central role of superspreading (called hubs in graph theory) in the contact network (see Supplementary Materials and Methods for more details).

How plausible is the time delay of 17 days between the effective *R* and the contact index? Since we have used the *R* value based on seven day intervals and a four day average of the case numbers we expect a delay of 7+2 days between the actual onset of symptoms and report in the table we have used [18]. Thus the delay of 17 days is close to the expected one if one adds four to seven days of incubation time [3]. The remaining mismatch could result from inaccuracies in the imputation of data where the onset of symptoms is not known or where no symptoms were reported [18].

It should be noted that the degree distribution is only one graph-based factor that enters the epidemic threshold. A real network could also have, for example, clustering and degree correlations that may change the *R* value [14]. This may also explain why the CI-*R*-relation in our computations does not go through the origin and varies over a much wider relative range than the seven-day *R* value for the first wave in Germany. Further work is needed to clarify the impact of these graph properties.

In summary, we showed a strong association between the effective *R* and the GPS assessed contact behavior of the German population. We found a high correlation of *R* with the contact index CI, which accounts for a heterogeneous contact behavior of the population and superspreading behavior. The CI allows to quantitatively evaluate risks for certain behavior for instance by regional analysis (see Fig. 2 in the Supplementary Material). Hence, we propose to use the Contact Index as a tool for an early warning system, to assess the effectiveness of social distancing policies and for decision support of social distancing policy-making. Overall, GPS data and various metrics derived from it can provide a means to assess the effect of lockdown measures on contacts and their association with reductions in infection rates.

## Supporting information

Supp Mat

## Data Availability

na

## Acknowledgements

We thank J. Kurths, E. Kolarczyk and V. Belik for valuable discussions

## Ethics and data safety

The effective reproduction number *R* values in our analysis have been obtained from the RKI Nowcasting website (https://www.rki.de/DE/Content/InfAZ/N/Neuartiges_Coronavirus/Projekte_RKI/Nowcasting_Zahlen.xlsx?__blob=publicationFile). They are based on register routine data and are publicly available for secondary analyses. The contact data is collected via a Software Development Kit (SDK) developed for the primary purposes of assessing the quality of cell phone networks. The general terms and conditions of this data collection (see www.netcheck.de/datenschutz) also cover the use for any secondary data analysis through a broad consent.

