## Supplementary material for "Forecasting the SARS-CoV-2 effective reproduction number using bulk contact data from mobile phones": Supp Mat

### Distribution of degrees

Evaluating the number of contacts per device we find that the degree of nodes, i.e., the number of contacts of each person, is broadly distributed with a long tail before the lockdown (Fig. 1, blue line) while at later dates after the first wave, the distribution has a much shorter tail and is highly concentrated around few contacts (compare brown and green lines). Thus initially there are many individuals with large numbers of contacts who would be potential 'super-spreaders' (called hubs in network theory), but the lockdown clearly led to a reduction of the number of such individuals at later weeks. The same pattern is observed before and after the second wave and lockdown (red and purple dots).

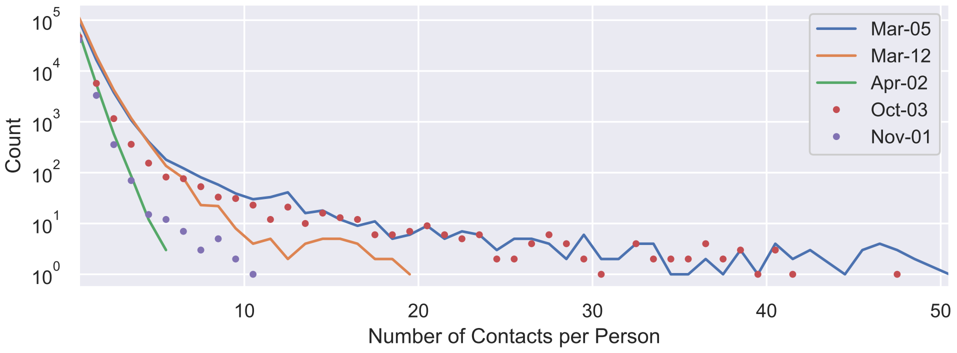


Figure 1: Histogram of number of contacts per person on three days before and after the first wave (solid lines) and two days before and during the second wave (dots): March 5, March 12 before official lockdown, March 19 (histogram cut off at 30 contacts) after ban of large gatherings and local regulations closing restaurant and other public venues. October 3 was a Saturday and also the national holiday with public gatherings. November 1 was one day before the second lockdown.

#### Local mapping of CI

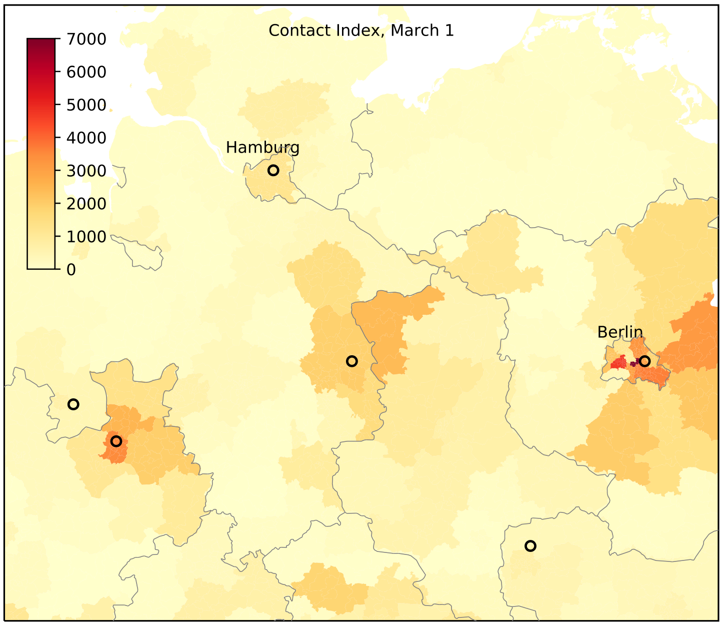


Figure 2: The CI can be locally calculated by only evaluating cell phones located mostly in a specific county (while keeping all contacts, as well outside of the county, of that cell phone for the calculation). The map shows the CI on March 1, 2020, a Sunday. Several soccer matches were hold on that day. The circles show cities that took part in the games from the two top tiers.

### Theory and Methods

In this Appendix, we describe our specially designed algorithm that identifies "contacts" from the traces based on the following rationale: If GPS pings arrive from two distinct cell phones that are close in space and time, then we denote this event as a "contact" and use it as a proxy for a human physical contact.

Our data allows estimating the number of real-world contacts for the entire population of Germany. However, a large part of these real contacts is missing from our cell phone sampling for two main reasons: (A) We only cover a fraction of devices. (B) We only cover times when the cell phone is sending a ping. In more detail regarding (A), we cover about 800,000 GPS-enabled devices per day, so that the majority of contacts for an individual goes undetected. As there are about 83 million persons in Germany, we can expect to cover about 1% of persons. Regarding (B), a typical cellphone sends about 200 pings per day. In order to cover the entire day, one ping every two minutes i.e. 720 pings per day are needed. Thus, only about 28% of the time of the day is covered for the average device. Assuming for simplification that the time of pings are independent for different devices, a lower bound estimate of the probability that a contact between two devices was observed is $0.28 \times 0.28 \approx 0.1$. So, according to this rough calculation, we can expect to track at least $0.01^2 \times 0.1 = 0.001\%$ of all contacts between any two persons living in Germany.

The average number of cell phones registered during a day was about 800,000. Per day we find between 20,000 and 160,000 devices that had at least one match. The total number of matched pairs varied between 150,000 (before lockdown) and 12,000 (during lockdown) per day.

#### Sampling of nodes

We now describe more formally how measures of the sampled network, such as mean contacts and second moment of contacts, relate to the respective measures of the original full contact network for all cell phones. We focus on sampling of devices described in restriction (A) in section ??, and ignore restriction (B) for simplification, since it is similar to the sampling of nodes in restriction (A) and would just require a re-scaling of the parameter $p$ in equation (14).

In the following, let $G$ denote the full network or graph of all cell phones and let $M$ denote the maximal degree of a node in $G$. As a reminder, the degree of a node (i.e. person) equals the number of contacts of this person. Following Zhang et al. [7] we let $N$ denote the vector containing the degree counts of the nodes (an alternative way of deriving our results is based on the Horvitz-Thompson estimate [2, 3]). $N$ has length $M+1$ and the $k$-th entry of $N$ contains the number of nodes that have degree $k$, i.e. the number of devices that have $k$ contacts. Thus $N$ contains the counts of the number of cell phones having $k$ links (contacts) to other cell phones.

In the sampling of phones according to (A), we assume that each phone is sampled from $G$ with the same probability $p$, resulting in the sampled graph $G^*$. This situation is also described as induced network sampling in network theory [7]. The induced network $G^*$ includes all sampled nodes as well as all links from $G$ that connect the sampled nodes in $G^*$.

The vector of the expected values of the degree counts of the sampled network, $N^*$, is $E(N^*) = PN$, Here, $P$ is a matrix of entries $P(k, k')$ that describe the probability that a node of degree $k'$ in $G$ is selected and has degree $k$ in $G^*$. For induced sampling, $P$ is:

$$P_{\text{ind}}(k, k') = \begin{cases} \binom{k'}{k} p^{k+1}(1-p)^{k'-k} & \text{for } 0 \le k \le k' \le M, \\ 0 & \text{for } 0 \le k' < k \le M. \end{cases} \tag{1}$$

Thus the $k$-th entry $E(N^*(k)) = \sum_{k',k \le k'} N(k') \binom{k'}{k} p^{k+1}(1-p)^{k'-k}$.

In the following we assume that the particular sampling given by our mobile phone records gives rise to a $N^*_{\text{ind}}$, which can be approximated by $E(N^*)$ for large networks, from which we can calculate the degree moments for the original network.

#### Derivation of the contact index $C$

Let $\langle k \rangle$ denote the mean degree of nodes in $G$: $\langle k \rangle = \sum_{k=0}^{M} kN(k) / (\sum_{k=0}^{M} N(k))$. We first show that the mean $\langle k \rangle_{\text{ind}}$ of the sampled graph is linearly related to the mean of the original graph:

$$\begin{aligned} \langle k \rangle_{\text{ind}} &\approx \frac{\sum_{k=0}^{M^*} kE(N^*(k))}{\sum_{k=0}^{M^*} E(N^*(k))} & (2) \\ &= \frac{\sum_{k,k',k \le k'} kN(k') \binom{k'}{k} p^{k+1}(1-p)^{k'-k}}{\sum_{k,k',k \le k'} N(k') \binom{k'}{k} p^{k+1}(1-p)^{k'-k}} & (3) \\ &= \frac{p \sum_{k'} N(k') \sum_{k,k \le k'} k \binom{k'}{k} p^{k}(1-p)^{k'-k}}{p \sum_{k'} N(k') \sum_{k,k \le k'} \binom{k'}{k} p^{k}(1-p)^{k'-k}} & (4) \\ &= \frac{p^2 \sum_{k'} k'N(k')}{p \sum_{k'} N(k')} & (5) \\ &= p\langle k \rangle & (6) \end{aligned}$$

The equality of (4) and (5) follows since $\sum_{k,k \le k'} k \binom{k'}{k} p^k (1-p)^{k'-k}$ is the mean value of the binomial distribution $B(k', p)$ which equals $k'p$, and $\sum_{k,k \le k'} \binom{k'}{k} p^k (1-p)^{k'-k}$ is the sum of all probabilities in $B(k', p)$ which is 1. Similarly, we find for the second moment:

$$\langle k^2 \rangle_{\text{ind}} \approx \frac{\sum_k k^2 E(N^*(k))}{\sum_k E(N^*(k))} \tag{7}$$

$$= \frac{\sum_{k,k',k \leq k'} k^2 N(k') \binom{k'}{k} p^{k+1} (1-p)^{k'-k}}{\sum_{k,k',k \leq k'} N(k') \binom{k'}{k} p^{k+1} (1-p)^{k'-k}} \tag{8}$$

$$= \frac{p \sum_{k'} N(k') \sum_{k,k \leq k'} k^2 \binom{k'}{k} p^k (1-p)^{k'-k}}{p \sum_{k'} N(k') \sum_{k,k \leq k'} \binom{k'}{k} p^k (1-p)^{k'-k}} \tag{9}$$

$$= \frac{p \sum_{k'} (k'(k'-1)p^2 + k'p) N(k')}{p \sum_{k'} N(k')} \tag{10}$$

$$= p^2 \langle k^2 \rangle - (p^2 - p) \langle k \rangle \tag{11}$$

Here, (7) is the definition of the second moment, (10) follows from (9) since the second moment for the binomial distribution $B(k', p)$ is $p^2 k'^2 + k'(p - p^2)$ and $\sum_{k,k \leq k'} \binom{k'}{k} p^k (1 - p)^{k'-k} = 1$, and (11) follows from (10) because of the definitions of the first and second moments of $N(k')$. Finally, we describe how the ratio $\langle k^2 \rangle / \langle k \rangle$ of the original graph can be obtained from the sampled graph via $\langle k \rangle_{\text{ind}}$ and $\langle k^2 \rangle_{\text{ind}}$:

$$\frac{\langle k^2 \rangle}{\langle k \rangle} \approx \frac{\frac{1}{p^2} \left( \langle k^2 \rangle_{\text{ind}} - (p - p^2) \langle k \rangle \right)}{\langle k \rangle} \tag{12}$$

$$= \frac{\langle k^2 \rangle_{\text{ind}}}{p \langle k \rangle_{\text{ind}}} - \left( \frac{1}{p} - 1 \right) \tag{13}$$

$$= \frac{1}{p} \left( \frac{\langle k^2 \rangle_{\text{ind}}}{\langle k \rangle_{\text{ind}}} - 1 \right) + 1. \tag{14}$$

This ratio $\langle k^2 \rangle / \langle k \rangle$ is of interest, since it describes the growth rate of an infection phase in an uncorrelated network [5]. Since $\langle k^2 \rangle_{\text{ind}}$ is larger or equal to $\langle k \rangle_{\text{ind}}$, (14) is non-negative and since $p$ is small in our sampling, we can ignore the addition of the constant 1. Thus we define the contact index $C$ as

$$\text{CI} := \frac{N_{\text{tot}}}{N_{\text{obs}}} \left( \frac{\langle k^2 \rangle_{\text{ind}}}{\langle k \rangle_{\text{ind}}} - 1 \right),$$

where $p$ has been replaced by the ratio $N_{\text{obs}}/N_{\text{tot}}$, where $N_{\text{obs}}$ is the number of devices observed during a day and $N_{\text{tot}}$ is the total number of devices/consumers in the considered area.

#### Effective R calculation

The effective reproduction number $R$ values in our analysis have been obtained from the RKI Nowcasting website [1]. For a given day $d$, $R$ is calculated as the ratio of the sums of infections for days $d$ to $d+6$ and $d-7$ to $d-1$ [6]. This number is then attributed to day $d$. For regional evaluations, confirmed cases are counted by the district where the individual has their home address.

#### Statistical analysis

In the analysis, we estimate the contact index from a sample of nodes from the full network of cell phones in Germany. We present descriptive statistics and temporal trends of the number of contacts as well as of the contact index. Finally, we investigate their association with infection rates assessed by $R$ by estimating their Pearson correlation coefficient. The Pearson correlations and their $p$-values were determined using Python's Scipy package, version 1.3.1.

#### GPS location data

The investigation relies on GPS location history data that is collected via a Software Development Kit (SDK) developed for the primary purposes of assessing the quality of cell phone networks. Cell phone data is collected by the SDK implemented in more than one million cell phones in Germany. Per day data was received from 1.15 to 1.4 million cell phones during March to July 2020. The legal conditions for the processing of the data were described in a report by A. Böken on May 11, 2020. Data records are anonymous. In a first step the number of contacts for each device is determined so that no positional information is retained. Then the data is aggregated by the number of devices that have a certain number of contacts. Only these aggregated numbers are used for further analysis.

#### Probability of contact detection and real distance between individuals

The epidemiology of SARS-CoV-2 indicates that most infections are caused by close contacts through respiratory droplet transmission with a short range, e.g. 1.5 to 2 m [4] A feasible algorithm for contact detection from GPS data needs to have acceptable accuracy, while still providing efficacy in regards to computation time. Since GPS pings of several million individuals are assessed over a longer time span, a vast amount of data needs to be analyzed. Therefore, we map GPS positions to an 8m x 8m grid and assign a time stamp with a resolution of 2 minutes. Each tile within the grid has a unique identification number (tile ID). If entries have identical tile IDs and time stamps (which were

[1] https://www.rki.de/DE/Content/InfAZ/N/Neuartiges_Coronavirus/Projekte_RKI/Nowcasting_Zahlen.xlsx?__blob=publicationFile

rounded to two minutes), this is accounted as a contact. Contact detection might fail in dependence of the choice of grid placement, if GPS positions get mapped to different tiles even though they are in close proximity. This effect becomes more likely with increasing distance between GPS coordinates.

For evaluation of this algorithm, we used a numerically approach: we set a point $p_1$ to the origin coordinates and a point $p_2$ to random coordinates and distance $d$ to $p_1$. We then create a tile with random positions which includes $p_1$. If $p_2$ is also within the tile, the contact with distance $d$ was successfully detected. By repeating this algorithm, a curve for the probability of contact detection in relation to distances between GPS positions can be calculated (Fig. 3A). To calculate a similar graph for the real positions between individuals, the inaccuracy of GPS has to be taken into consideration. In order to do this, we randomly shifted $p_1$ and $p_2$ according to the known inaccuracy of GPS (mean error 0.95 m +- 1.05 m standard deviation [1] The result can be seen in Fig. 3B.

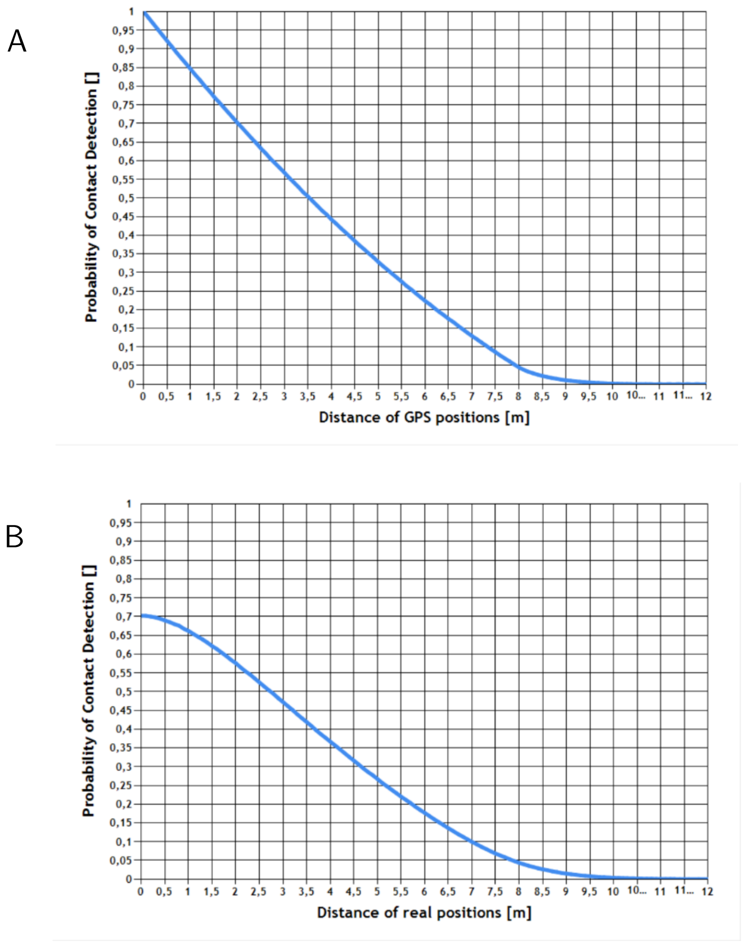


Figure 3: Probability of contact detection. See text for a details.
